# Serum asymmetric dimethyl arginine level correlates with the progression and prognosis of amyotrophic lateral sclerosis

**DOI:** 10.1101/2021.07.01.21259228

**Authors:** Kensuke Ikenaka, Yasuhiro Maeda, Yuji Hotta, Seiichi Nagano, Keita Kakuda, Harutsugu Tatebe, Naoki Atsuta, Daisuke Ito, Cesar Aguirre, Yasuyoshi Kimura, Kousuke Baba, Masahisa Katsuno, Takahiko Tokuda, Kazunori Kimura, Gen Sobue, Hideki Mochizuki

**Author notes:** **Corresponding Author:** Kensuke Ikenaka, Department of Neurology, Osaka University Graduate School of Medicine, 2-2 Yamadaoka, Suita 565-0871, Japan.

## Abstract

**Objective:** To investigate the association between serum asymmetric dimethylarginine (ADMA) levels and the progression and prognosis of amyotrophic lateral sclerosis (ALS), and to compare cerebrospinal fluid (CSF) and serum ADMA levels with other biomarkers of ALS.

**Methods:** Serum ADMA levels of patients with sporadic ALS (n = 68) and disease control patients (n = 54) were measured using liquid chromatography-tandem mass spectrometry. Serum samples were obtained at the time of patient registration for diagnosis. Correlations of ADMA level and other markers (nitric oxide [NO] and neurofilament light chain [NFL] levels) were analyzed. Changes in the ALS Functional Rating Scale–Revised (ALSFRS-R) score from the onset of disease (ALSFRS-R preslope) was used to assess disease progression. Survival was evaluated using the Cox proportional hazards model and Kaplan–Meier analysis.

**Results:** The concentration of ADMA in CSF was substantially higher in patients with ALS than in disease controls. Serum ADMA level correlated with CSF ADMA level (*r* = 0.591, *p* < 0.0001), and was independently associated with the ALSFRS-R preslope (*r* = 0.505. *p* < 0.0001). Patients with higher serum ADMA levels had less favorable prognoses. CSF ADMA level significantly correlated with CSF NfL level (*r* = 0.456, *p* = 0.0002) but not with NO level (*r* = 0.194, *p* = 0.219).

**Conclusion:** ADMA level is an independent biomarker of ALS disease progression and prognosis, and reflects the degree of motor neuron degeneration. The increased ADMA level in ALS patients was not associated with the inhibition of NO production.

## Introduction

Amyotrophic lateral sclerosis (ALS) is a fatal neurodegenerative disease characterized by the selective loss of upper and lower motor neurons. Despite decades of research into the molecular mechanisms underlying the pathogenesis of ALS and the development of molecular targeting therapy, to date, only a few drugs have been shown to be effective for ALS. One of the difficulties in performing clinical trials for ALS patients is the large variation in the clinical courses of the patients^1 2^, as evaluated by survival time or the revised ALS Functional Rating Scale (ALSFRS-R). To obtain better patient stratification and more reliable measures for monitoring the therapeutic effects of potential treatments, it is imperative to develop biomarkers that accurately reflect the pathophysiology and predict the progression and prognosis of ALS.

We recently reported that protein arginine dimethylation is upregulated in the spinal cord of patients with amyotrophic lateral sclerosis (ALS), and that cerebrospinal fluid (CSF) levels of asymmetric dimethylarginine (ADMA) can be a biomarker of ALS disease progression and prognosis^3^. We hence hypothesized that increased CSF ADMA levels might reflect the hypermethylation of RNA-binding proteins in the spinal motor neurons and surrounding glial cells of ALS disease patients. Interestingly, recent reports have shown that RNA-binding proteins that are targets of arginine dimethylation demonstrate abnormal aggregation or mislocalization in the motor neurons of sporadic ALS (SALS) patients^4-7^, indicating the involvement of abnormal arginine dimethylation in the pathogenesis of ALS.

In this study, we analyzed the changes in serum ADMA levels in ALS patients and the correlation of ADMA level with neurofilament light chain (NfL) level, to understand how ADMA is associated with the pathology of ALS. We also analyzed the correlation between ADMA and nitric oxide (NO) levels in the CSF to determine whether increased ADMA affects ALS pathology through the suppression of NO synthesis. Moreover, towards clinical application, we analyzed the usefulness of serum ADMA level for predicting disease progression and prognosis.

## Materials and methods

### Patient registry and follow-up

Patients who were diagnosed as having ALS at Osaka University between July 2016 and August 2020 were prospectively enrolled. The data were collected from the patients agreed to participate in the Osaka University Longitudinal Biomarker Study for Neuromuscular Diseases. In total, 68 ALS patients with definite, probable, probable laboratory-supported, or possible ALS according to the revised El Escorial criteria were included. The included patients were registered in the Osaka University Longitudinal Biomarker Study for Neuromuscular Diseases with written informed consent. The clinical scores listed below were obtained for the diagnosis of ALS. Muscle strength was manually tested and scored using the scale of the Medical Research Council (manual muscle testing [MMT]). Disease onset was defined as when the patients became initially aware of muscle weakness or the impairment of swallowing, speech, or respiration. The Japanese version of the ALS Functional Rating Scale-Revised (ALSFRS-R) ^2^ was used as a scale to evaluate ADL. The reliability of the Japanese version of the ALSFRS-R has been confirmed previously^8^. To evaluate the functional decline in the ALSFRS-R, we calculated the slope, defined as (decrease in the value within a duration)/duration. The preslope was used to evaluate the decline in the ALSFRS-R from the time of onset to diagnosis (registration), and was calculated as ALSFRS-R preslope = (ALSFRS-R at registration − 48)/(duration from onset to diagnosis). The primary endpoint was defined as either the introduction of tracheostomy positive pressure ventilation (TPPV) or death of the patient, and the time a patient reached the primary endpoint was determined by telephone follow-up. TPPV-free survival was defined as survival in the TPPV cases.

### Clinical data of ALS and control patients

The average age at registration (ALS, 59.6 [46.9–72.3] years and controls, 62.0 [48.5–71.5] years) and sex ratio were not significantly different between patients with ALS and controls; sex ratio [male: female]: ALS, 41: 27 and controls, 32: 22). The average duration from onset to registration (months) in patients with ALS was 21.1 (5.12–37.1), the average ALSFRS-R at registration was 38.7 (32.5–44.9), and the average %forced vital capacity (FVC) at registration was 84.9% (63.0%––106.8%), which are consistent with previous studies^2,9-11^. At the end of this study, 23 patients reached the primary endpoint, and the average duration from registration to the primary endpoint was 18.9 months. Regarding disease form (initial symptoms), 13 patients showed the bulbar form, and 55 patients showed the spinal form. The disease controls included 54 patients with Parkinson disease (n = 10), Parkinson syndrome (n = 9), polyneuropathy (n = 7), multiple sclerosis (n = 6), myositis (n = 6), brain infarction (n = 5), dystonia (n = 3), and others (n = 8).

### Measurement of arginine analogs

Arginine, NG, and NG-dimethyl L-arginine (ADMA) were used to construct standard curves (Enzo Life Sciences). ADMA-d6 was prepared according to the method of Kennedy et al.^12^. ADMA was measured using a high-performance liquid chromatography-tandem mass spectrometry system (Quattro Premier XE Mass Spectrometer; Waters Corporation, Milford, MA, USA). A 5-μL sample solution that was deproteinated by methanol was injected into an Intrada Amino Acid column (2 × 50 mm; Imtakt, Kyoto, Japan) at 40 °C. Chromatography was performed at a flow rate of 0.6 mL/min using a step gradient alternating between a mixture of acetonitrile: tetrahydrofuran: 25 mmol/L aqueous ammonium formate: formic acid (9: 25: 16: 0.3) and a mixture of 100 mmol/L aqueous ammonium formate: acetonitrile (80: 20). ADMA was analyzed using the multiple reaction monitoring mode of tandem mass spectrometry in positive ion mode. The cone voltage was 22–25 V, collision energy was 13–22, and transitions were m/z 203 → 46 for ADMA.

### Quantification of plasma and CSF NfL concentrations

The concentrations of plasma and CSF NfL were quantified as previously described^13, 14^,, using Simoa NF-light Advantage Kit and a Simoa HD-1 analyzer according to the manufacturer’s protocol (Quanterix, Lexington, MA, USA). All samples were analyzed in duplicate.

### Measurement of NO concentration

The nitrite concentration in the CSF was measured using the nitrite assay kit following the manufacturer’s instructions (BioVision, Milpitas, CA, USA). Absorbance at 540 nm was measured using a microplate reader.

### Statistical analysis

Pearson’s correlation analysis was performed to analyze the correlation between factors. Survival time was defined as the time from disease onset to death or the introduction of TPPV. The Kaplan-Meier method was used to estimate survival curves, and the survival curves of the two groups were compared using the logrank test. The Cox proportional hazard model, which included the ALSFRS-R slope at registration and serum ADMA level (ng/mL), was applied to analyze the effects of these variables on survival time. The hazard ratio (HR) and 95% confidence interval (CI) were estimated. Multivariate regression analyses with stepwise variable selection (alpha = 0.05 for inclusion, and alpha = 0.10 for exclusion) was also performed to analyze the effect of ADMA on ALSFRS-R. Statistical Package for the Social Sciences 23.0J software (IBM Japan, Tokyo, Japan) was used to perform statistical analyses.

### Ethics statement

This study was conducted in accordance with the Declaration of Helsinki and the Ethical Guidelines for Medical and Health Research Involving Human Subjects endorsed by the Japanese government. The Ethics Committee of Osaka University Graduate School of Medicine approved the study (Approval number; 19089-3). All the patients, including disease controls, were informed about this study and written consent have been obtained.

## Results

### Serum ADMA level is increased in ALS patients and correlates with disease progression

We first compared ADMA levels in the serum and CSF, and found that serum ADMA level significantly correlated with CSF level (Figure 1A, *r* = 0.591, *p* < 0.032). Similarly to the increase in CSF ADMA level that we found in our previous study, serum ADMA level was also significantly higher in ALS patients than in disease controls (Figure 1B, *p* = 0.002). Serum ADMA level more strongly correlated with disease progression (ALSFRS-R preslope) (Figure 1C, *r* = 0.505. *p* < 0.0001) than disease severity at each timepoint (ALSFRS-R) (Figure 1D, *r* = −0.261, *p* = 0.032). More importantly, multivariate linear regression analysis demonstrated that among serum ADMA, age, sex, MMT, %FVC, and ALSFRS-R, only serum ADMA was an independent factor that was associated with disease progression (Table 1).

**Table 1.**
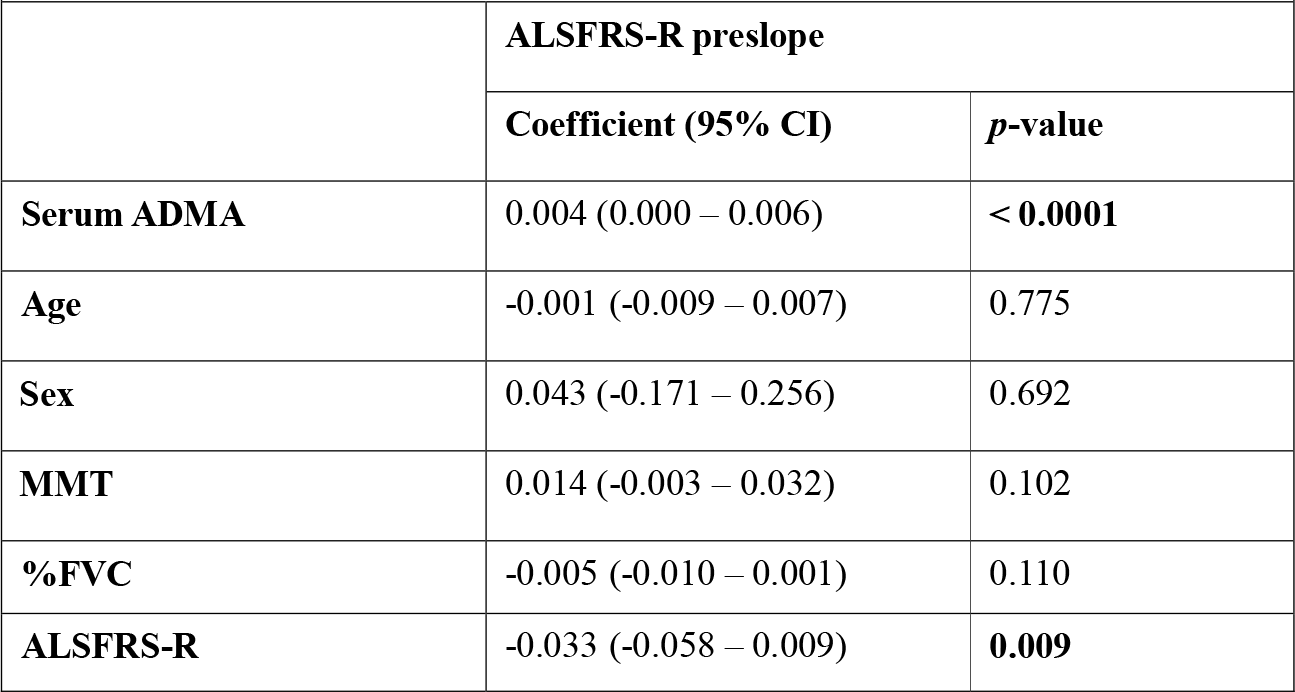
Multivariate linear regression analysis with stepwise variable selection; comparison among serum ADMA and clinical ALS parameters.

|  |  |  |
| --- | --- | --- |
|  | **ALSFRS-R preslope** |  |
|  | **Coefficient (95% CI)** | ***p*-value** |
| **Serum ADMA** | 0.004 (0.000 – 0.006) | **< 0.0001** |
| **Age** | -0.001 (-0.009 – 0.007) | 0.775 |
| **Sex** | 0.043 (-0.171 – 0.256) | 0.692 |
| **MMT** | 0.014 (-0.003 – 0.032) | 0.102 |
| **%FVC** | -0.005 (-0.010 – 0.001) | 0.110 |
| **ALSFRS-R** | -0.033 (-0.058 – 0.009) | **0.009** |

**Figure 1.**
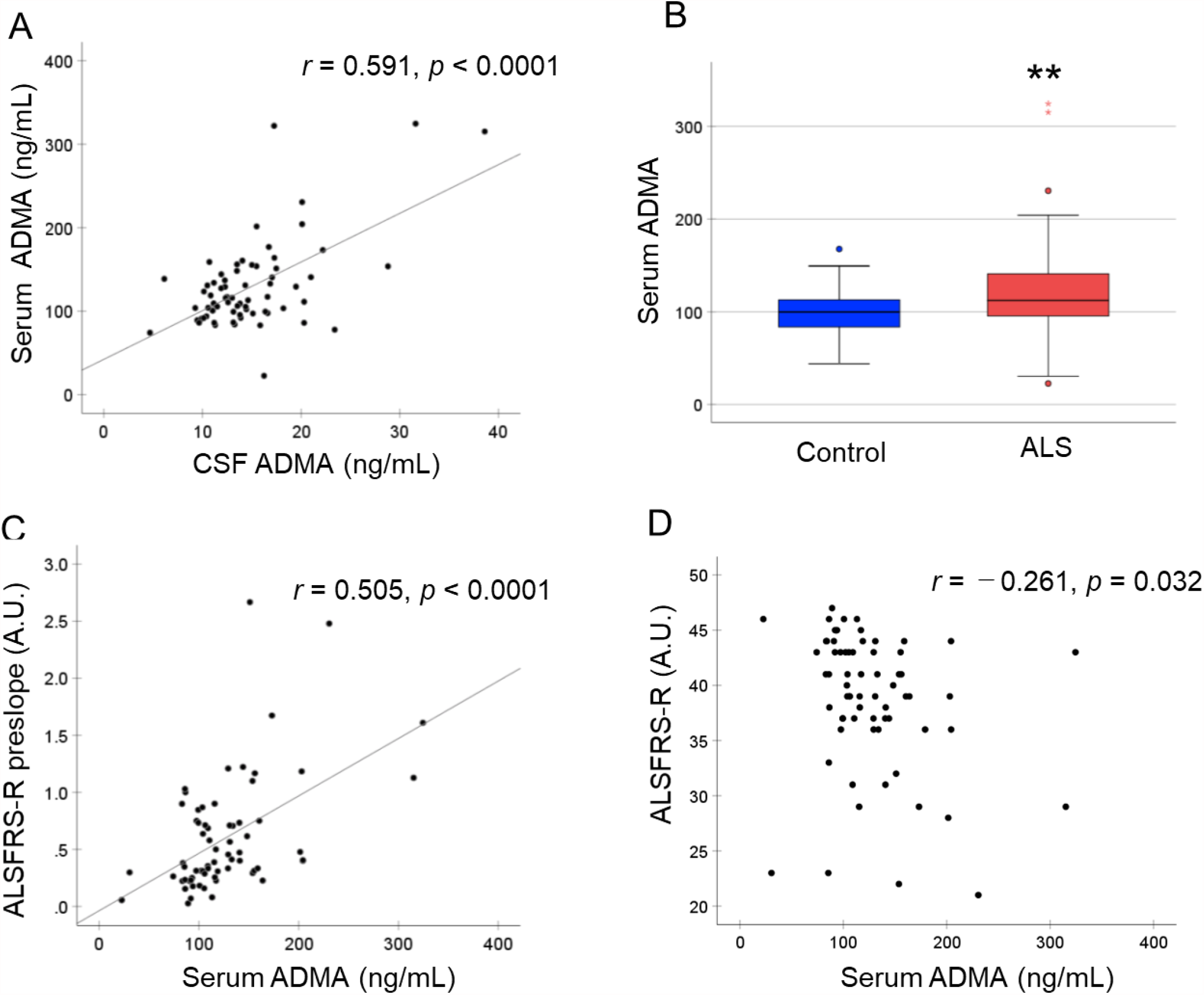
Increased serum ADMA level in ALS patients and its association with disease progression. (A) Correlation between CSF and serum ADMA levels (n = 68). (B) ADMA concentrations of control patients (n = 54) and ALS patients (n = 68). \*\**p* < 0.01, Student *t*-test. (C) Correlation of serum ADMA concentrations and disease progression scores (ALSFRS-R preslope) or disease severity score (ALSFRS-R) (n = 68 for both analyses).

### Associations between serum ADMA levels and other biomarkers

Moreover, we measured plasma NfL levels to analyze the degree of neurodegeneration in patients, and found that ADMA level strongly correlated with NfL level (Figure 2A, *r* = 0.430, *p* = 0.002). Interestingly, ADMA level did not correlate with NO level (Figure 2B, *r* = 0.270, *p* = 0.080), indicating that the increase in ADMA level was independent of NO dysregulation. Multivariate linear regression analysis demonstrated that among serum ADMA, creatinine, albumin, NO, and plasma NfL level, serum ADMA level was independently associated with disease progression (Table 2).

**Table 2.** Multivariate linear regression analysis with stepwise variable selection; comparison among serum ADMA and other biomarkers.

|  | ALSFRS-R preslope |  |
| --- | --- | --- |
|  | Coefficient (95% CI) | *p*-value |
| **Serum ADMA** | 0.006 (0.001–0.011) | **0.024** |
| **Creatinine** | -0.536 (-1.255 – 0.183) | 0.139 |
| **Albumin** | -0.30 (-0.429 – 3.69) | 0.880 |
| **Nitric oxide** | -0.003 (-0.048 – 0.041) | 0.878 |
| **Plasma NfL** | 0.002 (0.000 – 0.004) | **0.045** |

**Figure 2.**
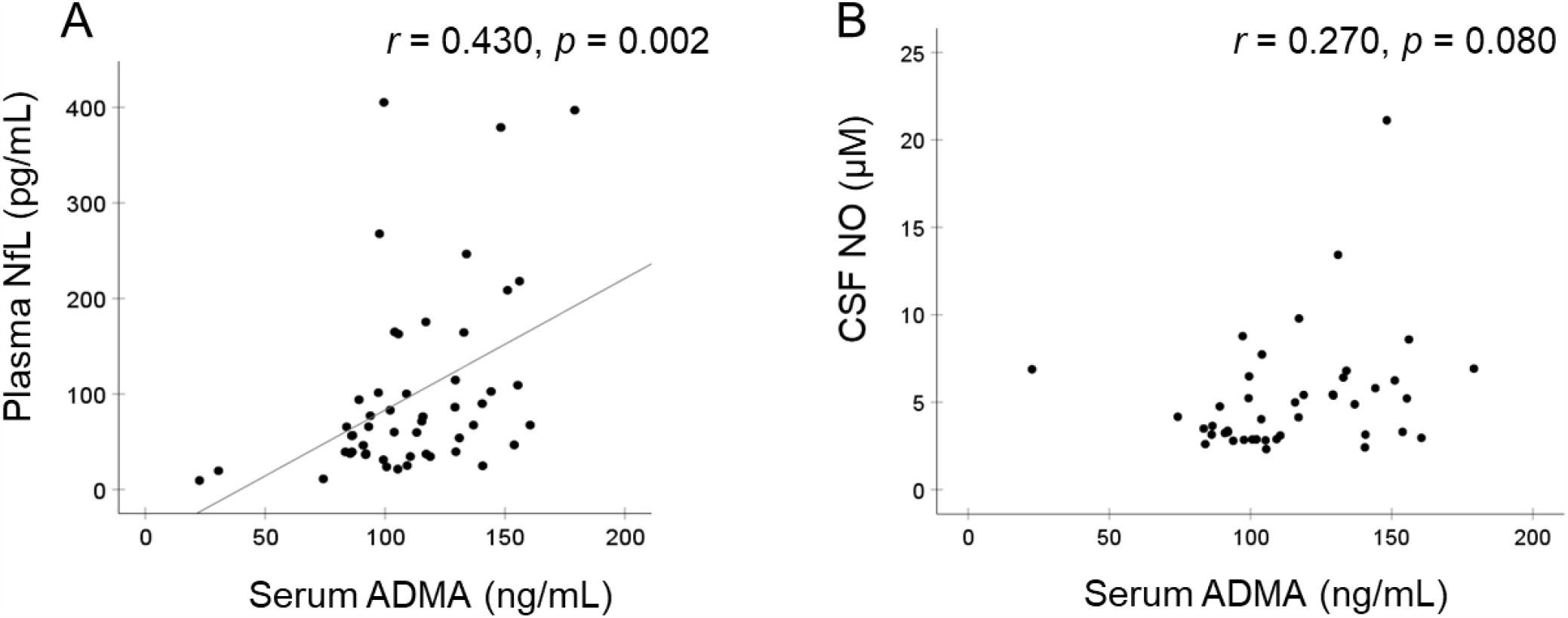
Comparison between serum ADMA level and plasma NfL or CSF NO level. (A) Correlation between serum ADMA concentration and plasma NfL level (n = 51). (B) Correlation between serum ADMA level and plasma NfL level (n = 43).

### Serum ADMA is a prognostic marker for ALS

Next, we performed multivariate Cox regression analysis of survival time, and found that serum ADMA level can predict the survival of patients independently of the ALSFRS-R preslope (Table 3).

**Table 3.** Multivariate Cox regression analysis of the survival of ALS patients, with adjustments of covariates.

|  | Primary endpoint |  |
| --- | --- | --- |
|  | HR (95% CI) | *p*-value |
| **Serum ADMA** | 1.008 (1.000 – 1.016) | **0.040** |
| **ALSFRS-R preslope** | 4.368 (1.771 – 10.771) | **0.001** |
| **n = 68**<br>**(23 subjects reached the**<br>**primary endpoint and 38**<br>**subjects were censored)** |  |  |

We then analyzed the optimal cutoff score of ADMA level for predicting the prognosis of ALS. When we used a cutoff of ADMA > 110.53 ng/mL, the HR was 4.289 (95% CI: 1.642–11.235, *p* = 0.003). We divided the registered patients into two categories using this cutoff score. Figure 3A shows the Kaplan–Meier curves for the primary endpoint of patients in the two categories. The difference between the curves was statistically significant by the logrank test (*p* = 0.001).

**Figure 3.**
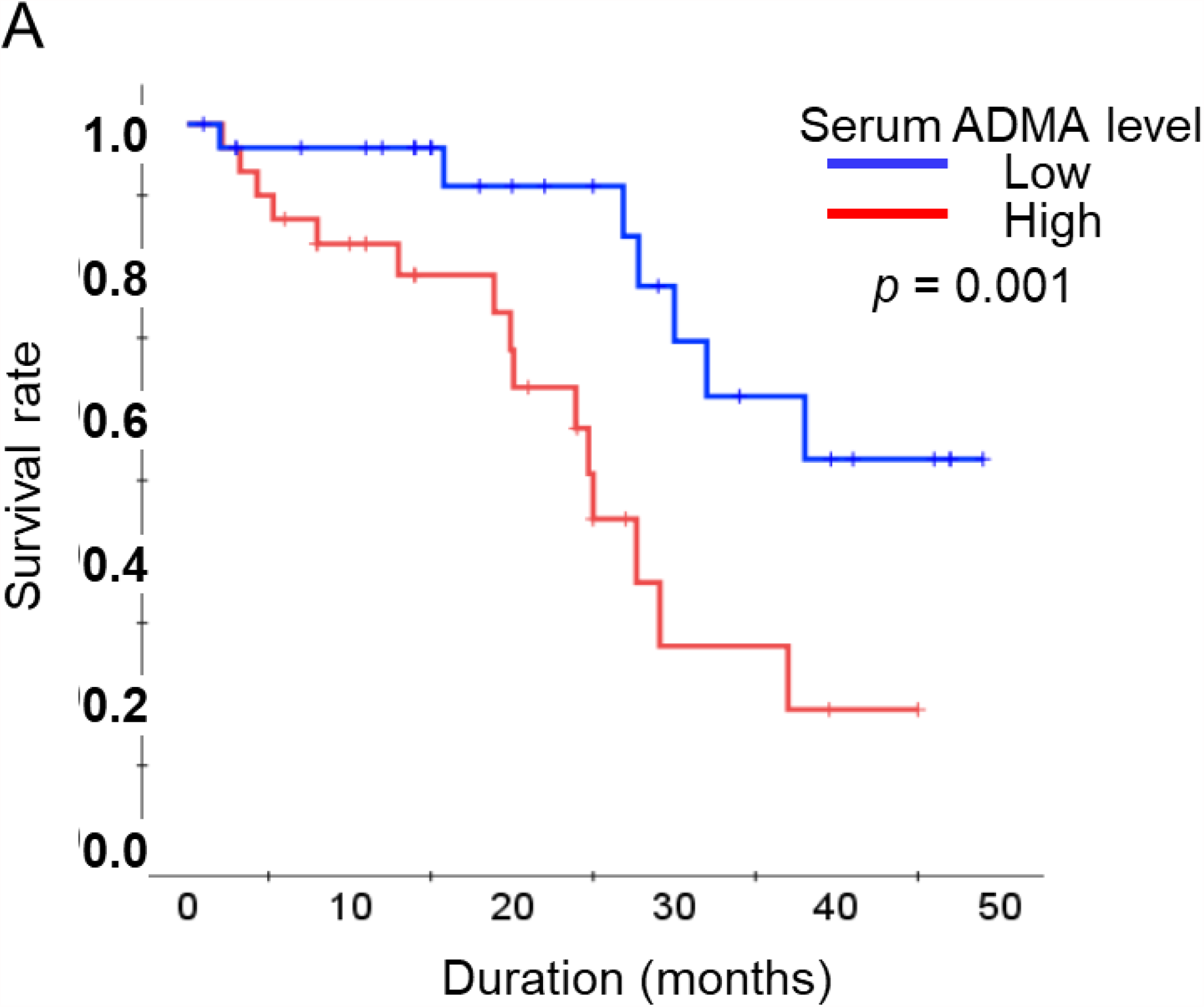
Serum ADMA level predicts the prognosis of ALS patients. (A) Kaplan-Meier curve according to serum ADMA level (low: < 110.53 ng/mL, blue line; high: > 110.54, red line). Kaplan-Meier curves for the primary endpoint were compared by the logrank test. There was a statistically significant difference between the curves (*p* = 0.001).

## Discussion

In this study, we showed that serum ADMA level is well correlated with CSF ADMA level and is useful for evaluating ALS disease progression and prognosis, similarly to what we previously reported for CSF ADMA^15^. Interestingly, ADMA level correlates more strongly with the ALSFRS-R slope than the ALSFRS-R. This suggests that unlike the decrease in creatinine level, which simply reflects decreased muscle volume and is hence just a consequence of ALS symptoms, the increase in ADMA level more accurately reflects the disease state of patients. Indeed, we showed that serum ADMA level correlates with plasma NfL level, which reflects axonal degeneration and is a pathological biomarkers for ALS^13,14,16-20^.

Here we showed that serum ADMA level is increased in ALS patients and is associated with disease progression score (ALSFRS-R slope) independently of several clinical scores and blood biomarkers, including plasma NfL. Serum ADMA level also predicts disease prognosis independently of the ALSFRS-R slope. We used the ALSFRS-R slope only as a coanalysis factor, because the number of patients who reached the primary endpoint (death or the use of an invasive respiratory machine) was only 23 among the 68 participants. Nevertheless, considering that the ALSFRS-R slope is one of the most reliable clinical predictors of prognosis at diagnosis, our data demonstrated that serum ADMA level may be a useful biomarker to predict the prognosis of ALS.

In our previous study, we mainly focused on the ADMA/L-arginine ratio, considering that ADMA competes with nitric oxide synthase for binding to L-arginine and inhibits the production of NO. In the present study, we directly measured ADMA instead of its ratio with L-arginine, because we found that ADMA level did not correlate with CSF NO. Our findings suggested that a high ADMA level may reflect ALS pathology independently of the insufficient production of NO, and may shed light on the importance of the hypermethylation of arginines within RNA-binding proteins in the pathogenesis of SALS. Moreover, considering that serum ADMA level is about ten times higher than that of CSF ADMA level, systemic hypermethylation could be the primary change and affects the pathology of the central nervous system in ALS patients.

This study has some limitations and bias. First, the number of patients who reached the primary endpoint was small, so the analysis of prognosis was statistically weak. Second, the pathomechanism by which an increased ADMA level affects disease progression remains unclear. Further studies are required to confirm that arginine hyperdemethylation is directly involved in the pathogenesis of ALS, and to determine whether ADMA can be a useful biomarker for clinical applications.

## Data Availability

not applicable

## COMPETING INTERESTS

The authors have no conflicts of interest to declare.

## FUNDING

This work was supported by the Kanae Foundation for the Promotion of Medical Science and Japan Agency for Medical Research and Development (grant no. JP21wm0425013).

## Author contributions

KI, NA, MK, HM, and GS designed the experiments. YM, YH, and KK performed the arginine analysis by liquid chromatography-tandem mass spectrometry. HT and TT performed the neurofilament light chain (NfL) measurements. KI, SN, YK, and KB collected the blood and cerebrospinal fluid (CSF) samples and clinical data. KI performed the statistical analysis. KI, NA, DI, CA, TT, MK, GS, and HM discussed the results and wrote the manuscript. All authors read and approved the final manuscript.

### Abbreviations

ADL: activities of daily living
ADMA: asymmetric dimethyl arginine
ALS: amyotrophic lateral sclerosis
ALSFRS-R: ALS Functional Rating Scale-Revised
CI: confidence interval
CK: creatinine kinase
CSF: cerebrospinal fluid
FVC: forced vital capacity
HR: hazard ratio
MMT: manual muscle testing
NfL: neurofilament light chain
TPPV: tracheostomy positive pressure ventilation

## Notes

### Competing Interest Statement

The authors have declared no competing interest.

